# ACE inhibition and cardiometabolic risk factors, lung *ACE2* and *TMPRSS2* gene expression, and plasma ACE2 levels: a Mendelian randomization study

**DOI:** 10.1101/2020.04.10.20059121

**Authors:** Dipender Gill, Marios Arvanitis, Paul Carter, Ana I. Hernández Cordero, Brian Jo, Ville Karhunen, Susanna C. Larsson, Xuan Li, Sam M. Lockhart, Amy Mason, Evanthia Pashos, Ashis Saha, Vanessa Y. Tan, Verena Zuber, Yohan Bossé, Sarah Fahle, Ke Hao, Tao Jiang, Philippe Joubert, Alan C. Lunt, Willem Hendrik Ouwehand, David J. Roberts, Wim Timens, Maarten van den Berge, Nicholas A. Watkins, Alexis Battle, Adam S. Butterworth, John Danesh, Barbara E. Engelhardt, James E. Peters, Don D. Sin, Stephen Burgess

## Abstract

**Objectives:** To use human genetic variants that proxy angiotensin-converting enzyme (ACE) inhibitor drug effects and cardiovascular risk factors to provide insight into how these exposures affect lung *ACE2* and *TMPRSS2* gene expression and circulating ACE2 levels.

**Design:** Two-sample Mendelian randomization (MR) analysis.

**Setting:** Summary-level genetic association data.

**Participants:** Participants were predominantly of European ancestry. Variants that proxy ACE inhibitor drug effects and cardiometabolic risk factors (body mass index, chronic obstructive pulmonary disease, lifetime smoking index, low-density lipoprotein cholesterol, systolic blood pressure and type 2 diabetes mellitus) were selected from publicly available genome-wide association study data (sample sizes ranging from 188,577 to 898,130 participants). Genetic association estimates for lung expression of *ACE2* and *TMPRSS2* were obtained from the Gene-Tissue Expression (GTEx) project (515 participants) and the Lung eQTL Consortium (1,038 participants). Genetic association estimates for circulating plasma ACE2 levels were obtained from the INTERVAL study (4,947 participants).

**Main outcomes and measures:** Lung *ACE2* and *TMPRSS2* expression and plasma ACE2 levels.

**Results:** There were no association of genetically proxied ACE inhibition with any of the outcomes considered here. There was evidence of a positive association of genetic liability to type 2 diabetes mellitus with lung *ACE2* gene expression in GTEx (p = 4×10^−4^) and with circulating plasma ACE2 levels in INTERVAL (p = 0.03), but not with lung *ACE2* expression in the Lung eQTL Consortium study (p = 0.68). There were no associations between genetically predicted levels of the other cardiometabolic traits with the outcomes.

**Conclusions:** This study does not provide evidence to support that ACE inhibitor antihypertensive drugs affect lung *ACE2* and *TMPRSS2* expression or plasma ACE2 levels. In the current COVID-19 pandemic, our findings do not support a change in ACE inhibitor medication use without clinical justification.

**Summary boxes:** *What is already known on this topic:*

- Severe acute respiratory syndrome coronavirus 2 (SARS-CoV-2) is responsible for the current coronavirus disease 2019 (COVID-19) pandemic.
- Serine protease TMPRSS2 is involved in priming the SARS-CoV-2 spike protein for cellular entry through the angiotensin-converting enzyme 2 (ACE2) receptor.
- Expression of *ACE2* and *TMPRSS2* in the lung epithelium might have implications for risk of SARS-CoV-2 infection and severity of COVID-19.

*What this study adds:*

- We used human genetic variants that proxy ACE inhibitor drug effects and cardiometabolic risk factors to provide insight into how these exposures affect lung *ACE2* and *TMPRSS2* expression and circulating ACE2 levels.
- Our findings do not support the hypothesis that ACE inhibitors have effects on *ACE2* expression.
- We found some support for an association of genetic liability to type 2 diabetes mellitus with higher lung *ACE2* expression and plasma ACE2 levels, but evidence was inconsistent across studies.

## Introduction

Severe acute respiratory syndrome coronavirus 2 (SARS-CoV-2) is responsible for the current coronavirus disease 2019 (COVID-19) pandemic (2). Serine protease TMPRSS2 is involved in priming the SARS-CoV-2 spike protein for cellular entry through the angiotensin-converting enzyme 2 (ACE2) receptor (3-6). COVID-19 patients most frequently present with respiratory tract infection symptoms and respiratory failure is the main cause of death (7-12). It follows that expression of *ACE2* and *TMPRSS2* in the lung epithelium might have implications for risk of SARS-CoV-2 infection and severity of COVID-19 (3, 13, 14).

Emerging evidence suggests that patients with underlying cardiometabolic risk factors and airway disease are more likely to suffer with severe COVID-19 (7-12). It has been speculated that the angiotensin-converting enzyme inhibitor (ACEi) and angiotensin receptor blocker (ARB) classes of antihypertensive medication that are more commonly prescribed in patients with cardiometabolic risk factors might affect expression of *ACE2*, and thus affect susceptibility to SARS-CoV-2 infection and severity of consequent COVID-19 (15-21). Although ACE and ACE2 are both dipeptidyl carboxydipeptidases, they have distinct physiological effects. ACE cleaves angiotensin I to angiotensin II, which consequently activates the angiotensin II receptor type 1 pathway resulting in vasoconstriction and inflammation. In contrast, ACE2 degrades angiotensin II to angiotensin 1-7 and angiotensin I to angiotensin 1-9. Angiotensin 1-9 activates the Mas receptor to have vasodilatory and anti-inflammatory effects. Animal studies have supported effects of ACEi and ARB drugs on ACE2 expression and activity (22-29), with mixed findings for associations of ACEi and ARB drug use with ACE2 activity and levels in human tissues also reported (30-32). It is important that any causal effects of these medications and cardiometabolic traits on *ACE2* expression be further investigated. Identification of a mechanistic basis by which such exposures affect risk and severity of COVID-19 could provide useful insight for disease prevention and treatment. This could be used to inform optimal medication use and strategies for shielding vulnerable individuals, as well as improving the evidence base for public health campaigns.

The Mendelian randomization approach uses genetic variants related to an exposure as instrumental variables for investigating the effect of that exposure on an outcome (33). Genetic variants are treated analogously to treatment allocation in a randomized controlled trial. Typically, molecular measurements such as gene expression or circulating protein levels are regarded in Mendelian randomization investigations as exposure variables. Here, following the work of Rao, Lau and So (34), we treat these molecular measurements as the outcomes in our investigation. The aim of this study was to apply Mendelian randomization to investigate whether *ACE2* and *TMPRSS2* gene expression in the lung and circulating levels of ACE2 in the plasma are associated with 1) genetic variants in the *ACE* gene region that can be considered as proxies for the effect of ACEi drugs, and 2) genetic variants related to cardiometabolic risk factors.

## Methods

### Genetic associations with exposure variables

Two different genetic instruments that proxy ACEi drug effects were considered. First, we selected 17 single-nucleotide polymorphisms (SNPs) in the *ACE* locus that were associated with serum ACE concentration in the Outcome Reduction with Initial Glargine INtervention (ORIGIN) trial and did not have strong pairwise correlation (r^2^ < 0.1) (35). Accounting for correlation, these variants explain 29.2% of the variance in serum ACE concentration. Secondly, we selected a single SNP, rs4291, located at the *ACE* locus that was associated with systolic blood pressure (SBP) at p = 9×10^−20^ in a study of 757,601 European-ancestry individuals (36, 37). Each blood pressure lowering allele of this SNP was associated with a 0.28mmHg reduction in SBP (36). There were no other SNPs at this locus that were associated with SBP at a genome-wide level of significance (p < 5×10^−8^) and did not have strong pairwise correlation (r^2^ < 0.1) with the index variant (36).

We further considered six cardiometabolic traits as exposure variables: body mass index (BMI), chronic obstructive pulmonary disease (COPD), lifetime smoking index, low-density lipoprotein cholesterol (LDL-C), SBP and type 2 diabetes mellitus (T2DM). These traits were chosen as they have been associated with prognosis of COVID-19 (7-12). Genetic association estimates for these exposures were obtained from the publicly available genome-wide association study (GWAS) summary data sources listed in Table 1. Genetic variants selected as instruments were SNPs associated with the corresponding trait at a genome-wide level of statistical significance (p < 5×10^−8^) and were uncorrelated (r^2^ < 0.001). Clumping of correlated variants was performed using the TwoSampleMR package in R (38).

**Table 1.** Sources for exposure trait genome-wide association study summary data.

| Trait | Sample size | Population ancestry | Number of variants | Variance explained (%) | Reference |
| --- | --- | --- | --- | --- | --- |
| Body mass index (BMI) | 806,834 | European | 546 | 5.7 | (64) |
| Chronic obstructive pulmonary disease (COPD) | 35,735 cases and 222,076 controls | Predominantly European | 82 | 7.0 | (65) |
| Lifetime smoking index | 462,690 | European | 126 | 0.36 | (66) |
| Low-density lipoprotein cholesterol (LDL-C) | 188,577 | European | 80 | 7.9 | (67) |
| Systolic blood pressure (SBP) | 318,417 | British | 192 | 2.9 | (68) |
| Type 2 diabetes mellitus (T2DM) | 74,124 cases and 824,006 controls | European | 202 | 16.3 | (69) |

### Genetic associations with outcome variables

Genetic associations with the expression of *ACE2* and *TMPRSS2* in lung tissue were obtained from two sources: 1) the Gene-Tissue Expression (GTEx) project (39), and 2) the Lung eQTL (expression quantitative trait loci) Consortium (40).

GTEx genetic association estimates were obtained in 515 individuals of predominantly European (85%) ancestry. Whole genome sequencing was performed by the Broad Institute’s Genomics Platform and only common variants (minor allele frequency > 0.05) were retained. Genome-wide eQTL analysis was performed for the expression of *ACE2* and *TMPRSS2* in primary tissue samples taken from the lung. Genetic associations with imputed variants across the autosomal chromosomes were adjusted for five principal components from the genotype data, 60 probabilistic estimation of expression residuals factors (41), sequencing platform (Illumina HiSeq 2000 or HiSeq X), sequencing protocol (polymerase chain reaction-based or free), and sex. For each gene, expression values between samples were normalized using the trimmed means of M-values method in EdgeR (42). Expression values were normalized across samples using an inverse normal transformation.

Lung eQTL Consortium genetic association estimates were obtained in 1,038 individuals of European ancestry (40). Tissue samples were obtained at three different institutions: University of British Columbia, Laval University, and University of Groningen. Genome-wide eQTL analysis was performed for the expression for *ACE2* using probe set 100134205_TGI_at and two probe sets for *TMPRSS2*, 100130004_TGI_at and 100157336_TGI_at (subsequently referred to as 1 and 2). All participants were genotyped using the Illumina Human 1M Duo BeadChip and the genotypes were imputed using the Haplotype Reference Consortium reference panel. Expression values were first standardized for age, sex, and smoking status using robust linear regression. Genetic associations were estimated in each cohort separately using a linear additive genetic model. The estimates were combined across cohorts using an inverse-variance weighted model with fixed-effects.

Genetic association estimates with circulating plasma ACE2 levels were obtained in a subcohort of 4,998 blood donors enrolled in the INTERVAL BioResource (1). Plasma ACE2 levels were measured using a multiplex proximity extension immunoassay (Cardiovascular 2 panel, Olink Bioscience, Uppsala, Sweden). A total of 4,947 samples passed quality control. The data were pre-processed using standard Olink workflows including applying median centring normalization across plates, where the median is centred to the overall median for all plates, followed by log2 transformation to provide normalized protein levels (NPX). NPX values were regressed on age, sex, plate, time from blood draw to processing (in days), and season. The residuals were then rank-inverse normalized. Genotype data was processed as described previously (43). Genome-wide pQTL analysis was performed by linear regression of the rank-inverse normalized residuals on genotype in SNPTEST (44), with the first three components of multi-dimensional scaling as covariates to adjust for ancestry.

### Mendelian randomization analyses

For the analysis investigating genetically proxied ACEi drug effects using the rs4291 variant, we report the genetic associations with lung *ACE2* and *TMPRSS2* expression and plasma ACE2 levels per blood pressure-lowering allele.

For all other Mendelian randomization analyses, estimates were obtained from the inverse-variance weighted method under a random-effects model (45). For the polygenic analyses based on the *ACE* gene locus, we accounted for correlation between variants (46). Heterogeneity between Mendelian randomization estimates from different genetic variants for the same exposure trait was expressed using the I^2^ statistic (47). For any identified associations at p< 0.05, the weighted median (48), MR-Egger (49) and contamination-mixture methods (50), which are more robust to the inclusion of pleiotropic variants, were performed as sensitivity analyses.

Mendelian randomization estimates represent the change in the outcome per one standard deviation increase in genetically predicted levels of the exposure for continuous exposure traits and per unit increase in the loge odds of the exposure for binary traits. All outcome measures were rank-based inverse-normal transformed, and so changes in the outcome measures are in standard deviation units.

### Ethical approval, data availability and reporting

The data used in this work are obtained from published studies that obtained relevant participant consent and ethical approval. All variants used as instruments and their genetic association estimates were selected from publicly available data sources, and are provided in Supplementary Tables 1-8. GWAS summary data for all the outcomes considered in this study are publicly available at http://dx.doi.org/10.6084/m9.figshare.12102681 (GTEx), http://dx.doi.org/10.6084/m9.figshare.12102711 (Lung eQTL Consortium), and http://dx.doi.org/10.6084/m9.figshare.12102777 (INTERVAL). The results from the analyses performed in this work are presented in the main manuscript or its supplementary files. This paper has been reported based on recommendations by the STROBE-MR Guidelines (Research Checklist) (51). The study protocol and details were not pre-registered.

### Patient and public involvement

There was no patient or public involvement in the design or reporting of this study.

## Results

Results for the analyses investigating genetically proxied ACEi drug effects are displayed in Figure 1. There was no evidence of associations with any of the outcome measures in the analyses considering the variants associated with serum ACE (Figure 1a), or the variant associated with SBP (Figure 1b).

**Figure 1.**
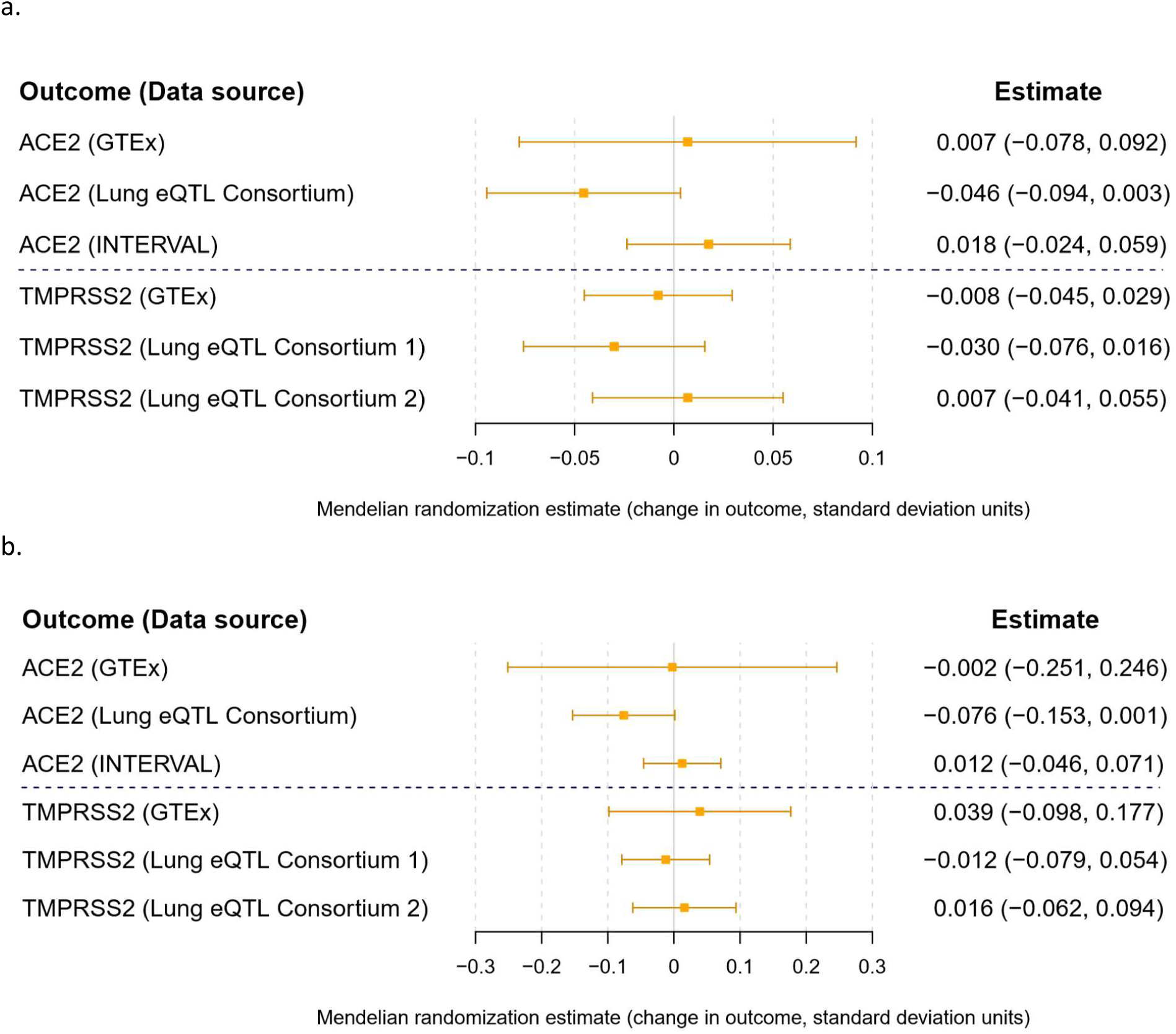
Genetic associations with *ACE2* and *TMPRSS2* gene expression in the lung (GTEx and Lung consortium) and circulating ACE2 protein levels in the plasma (INTERVAL): a) per one standard deviation increased ACE concentration conferred through variants at the *ACE* gene, and b) per blood pressure-lowering allele for the rs4291 variant in the *ACE* gene (bottom panel). The two sets of results for *TMPRSS2* expression in the Lung eQTL consortium refer to two separate probe sets for estimating gene expression.

The main inverse-variance weighted method Mendelian randomization results for the cardiometabolic risk factors are displayed in Figure 2 for lung *ACE2* expression and plasma ACE2 concentrations, and in Figure 3 for lung *TMPRSS2* expression. There was evidence of a positive association of genetic liability to T2DM with lung *ACE2* gene expression in GTEx (p = 4×10^−4^), and with circulating plasma ACE2 levels in INTERVAL (p = 0.03) (Supplementary Figure 1). Similar point estimates were obtained when performing the weighted median, MR-Egger and contamination-mixture Mendelian randomization sensitivity analyses that are more robust to the presence of pleiotropic variants, although the confidence intervals were wider (Supplementary Table 9). The MR-Egger method did not identify any evidence of directional pleiotropy biasing the analysis (Supplementary Table 9). There was no evidence of an association of genetic liability to T2DM with lung *ACE2* gene expression in the Lung eQTL Consortium (p = 0.68). There was no evidence of an association between genetically predicted levels of any of the other cardiometabolic traits with *ACE2* or *TMPRSS2* gene expression in GTEx or the Lung eQTL Consortium, or with circulating plasma ACE2 levels in INTERVAL.

**Figure 2.**
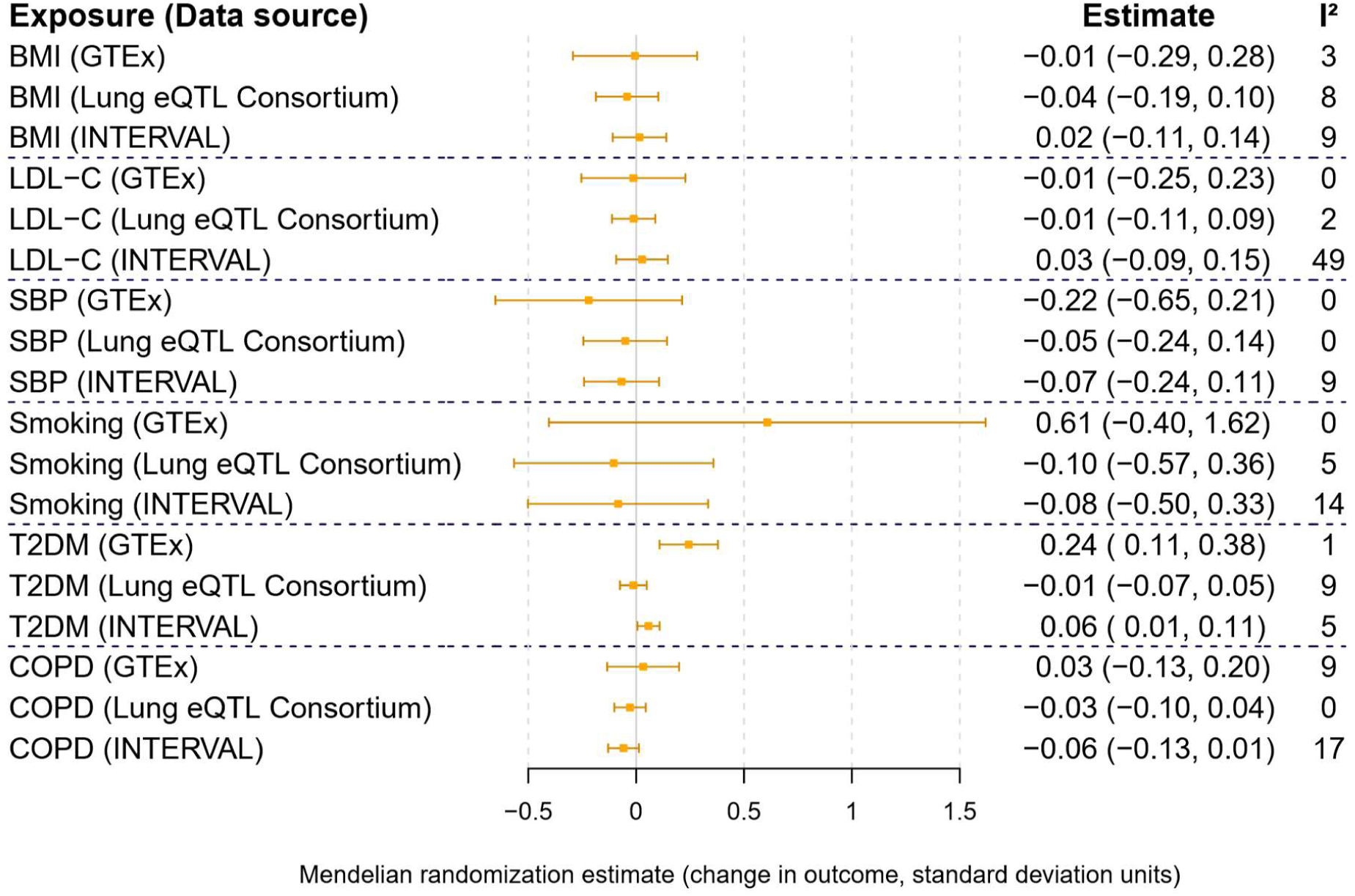
Mendelian randomization estimates for the change in *ACE2* gene expression in the lung (GTEx and Lung eQTL consortium) and circulating ACE2 protein levels in the plasma (INTERVAL) per unit increase in genetically predicted levels of the exposure.

**Figure 3.**
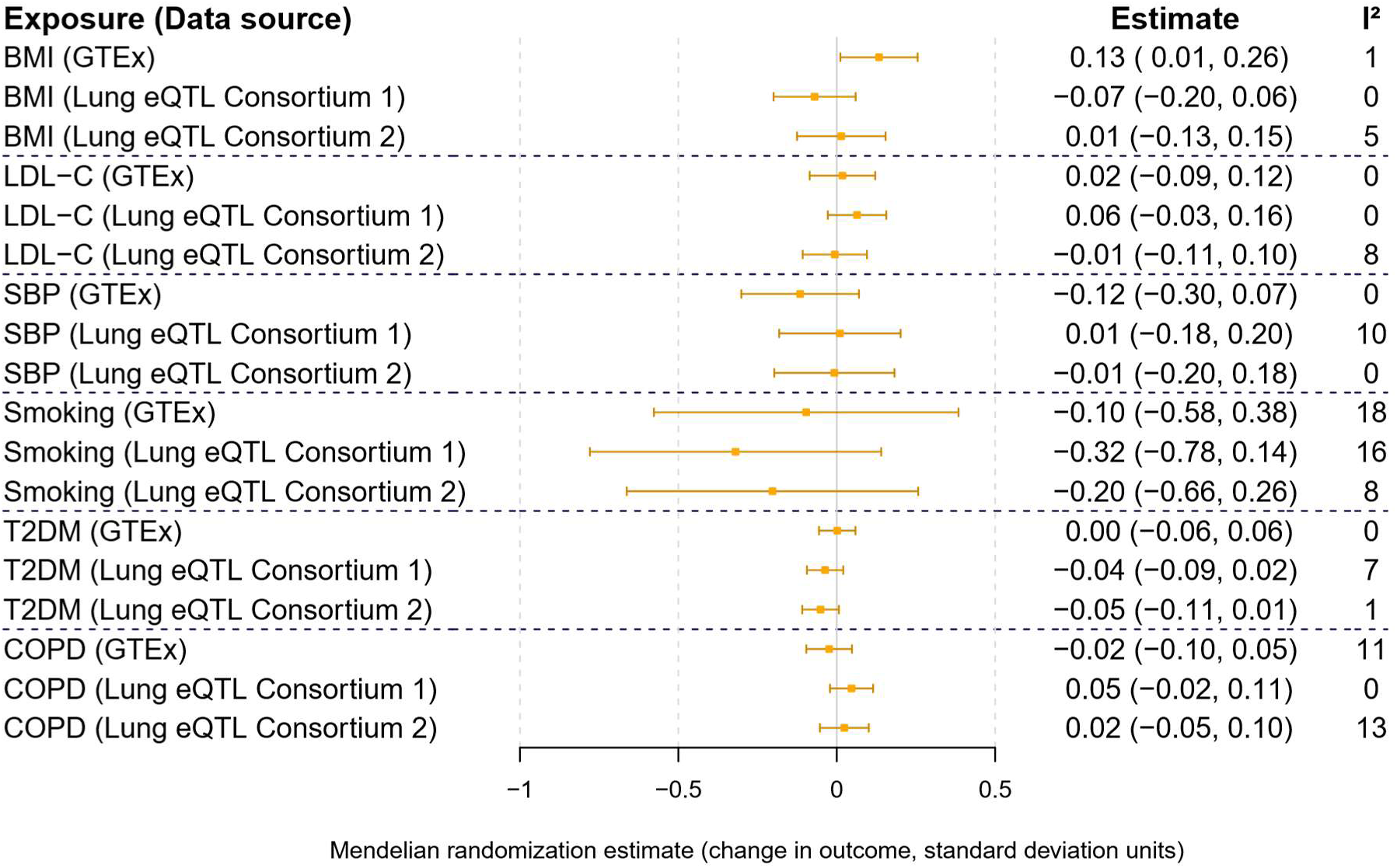
Mendelian randomization estimates for the change in *TMPRSS2* gene expression in the lung per unit increase in genetically predicted levels of the exposure. The two sets of results for the Lung eQTL consortium refer to two separate probe sets for estimating gene expression.

## Discussion

In the current COVID-19 pandemic, there is an urgent need to elucidate mechanisms underlying risk and severity of COVID-19, with a view to informing preventative and therapeutic strategies. In this study, we used human genetic variants that proxy ACEi drug effects and cardiometabolic risk factors to provide insight into how these exposures affect lung *ACE2* and *TMPRSS2* expression, and circulating ACE2 levels.

We did not find an association of genetically proxied ACE inhibition with lung *ACE2* and *TMPRSS2* expression or with circulating plasma levels of ACE2. These results therefore do not provide evidence to support that ACEi antihypertensive drugs affect risk or severity of COVID-19 through effects on ACE2 expression, as previously hypothesised (15-21). Previously identified changes in ACE2 expression in human tissues following ACEi treatment may not be applicable to the lung or circulating plasma levels (30, 31). Our findings support the stance of professional bodies for supporting continuation of ACEi and ARB antihypertensive drugs in patients with COVID-19 unless there is a clinical justification for stopping (52, 53). Indeed, appropriate use of these medications is of proven benefit (54, 55), and their abrupt interruption can also do considerable harm (56, 57). While there has also been speculation that ACEi and ARB antihypertensive drugs might reduce severity of COVID-19 (20, 58-60), with clinical trials to explore this currently planned (52), our findings are also consistent with guidance that patients should not start taking these drug classes unless clinically indicated (52).

Our results identified inconsistent support for an effect of liability to T2DM on lung *ACE2* expression and plasma ACE2 levels. An association of genetic liability to T2DM with lung *ACE2* expression in the GTEx project has previously been described (34). However, we identified no association of genetic liability to T2DM with lung *ACE2* expression in the Lung eQTL Consortium. This discrepancy may be attributable to the different populations considered in the GTEx project and the Lung eQTL Consortium (39, 40). While the Lung eQTL Consortium considered lung tissue from patients requiring resectional surgery, all samples in the GTEx project were taken from healthy tissue in deceased donors (39, 40). Taken together, our results do not provide consistent support for an effect of cardiometabolic traits on lung *ACE2* or *TMPRSS2* expression or plasma ACE2 levels. While the association between cardiometabolic traits and severity of COVID-19 could be attributable to alternative mechanisms (7-12), these risk factors can still be used to stratify patients in terms of their vulnerability. Similarly, while our current findings do not support a causal effect of COPD or smoking on lung *ACE2* expression, these factors may still be used to inform risk models for severe COVID-19 (61).

Our study has a number of strengths. We used genetic variants as instrumental variables for studying the effect of ACEi drugs and cardiometabolic risk factors, and were therefore able to investigate their causal effects on *ACE2* and *TMPRSS2* expression in the lung, and ACE levels in the plasma (33). For ACEi drugs effects, we used two complementary instrument selection strategies based on associations of variants at the *ACE* locus with circulating serum ACE levels and SBP respectively, and the consistent findings with both approaches add strength to our conclusions. Our Mendelian randomization approach is better able to overcome the confounding and reverse causation bias that can limit causal inferences from conventional epidemiological approaches (30, 62). Considering independent cohorts to assess lung expression of *ACE2* and *TMPRSS2* (39, 40), and plasma levels of ACE2 (43), we were able to explore consistency in our results and our conclusions are therefore less vulnerable to false positive findings.

Our study also has limitations. We only investigated *ACE2* and *TMPRSS2* expression in the lung and circulating levels of ACE2 in the plasma, and it may be that expression in other tissues is more relevant to risk and severity of COVID-19. Similarly, cellular ACE2 may have very different biological effects to circulating plasma ACE2. The GWAS analyses for lung *ACE2* expression and plasma ACE2 levels were all performed according to different protocols (39, 40, 43), and may therefore not be directly comparable. There was no available genetic instrument for the ARB antihypertensive drug class (36), and so we were not able to investigate this. The precision of our analyses was also limited, most notably for the lifetime smoking index results, which had widest confidence intervals. It may therefore be that our study was not sufficiently powered to exclude a clinically relevant effect for some exposures. The genetic variants that we used as instrumental variables may have pleiotropic effects where they affect the outcome through pathways independent of the exposure that they are proxying, and so bias the consequent Mendelian randomization estimates. While it is not possible to exclude this possibility, the relatively low heterogeneity detected between Mendelian randomization estimates produced by different variants, along with the consistency observed when performing analysis methods that are more robust to pleiotropy, suggests that this is unlikely to be a major source of bias (47).

In summary, this Mendelian randomization study does not identify consistent evidence to support that ACEi antihypertensive drugs or cardiometabolic traits affect lung expression of *ACE2* and *TMPRSS2*, or plasma ACE2 levels. These findings therefore do not support deviation from existing expert consensus guidelines for the management of hypertension in the face of the current COVID-19 pandemic (52). Efforts should be made by scientists and the news media to ensure that speculative stories with little evidential support are not propagated (63). While cardiometabolic risk factors can be used to stratify patients in terms of their vulnerability to COVID-19, our data do not provide consistent support that expression of *ACE2* or *TMPRSS2* represents causal mechanisms underlying these associations.

## Data Availability

The data used in this work are obtained from published studies that obtained relevant participant consent and ethical approval. All variants used as instruments and their genetic association estimates were selected from publicly available data sources, and are provided in Supplementary Tables 1-8. GWAS summary data for all the outcomes considered in this study are publicly available at http://dx.doi.org/10.6084/m9.figshare.12102681 (GTEx), http://dx.doi.org/10.6084/m9.figshare.12102711 (Lung eQTL Consortium), and http://dx.doi.org/10.6084/m9.figshare.12102777 (INTERVAL).

http://dx.doi.org/10.6084/m9.figshare.12102711

http://dx.doi.org/10.6084/m9.figshare.12102777

## Acknowledgements

We would like to thank those children and family members who allowed us time to work on this manuscript despite disruptions to schooling and daily routine. Participants in the INTERVAL BioResource were recruited with the active collaboration of NHS Blood and Transplant England (www.nhsbt.nhs.uk), which has supported field work and other elements of the trial. DNA extraction and genotyping was co-funded by the National Institute for Health Research (NIHR), the NIHR BioResource (http://bioresource.nihr.ac.uk) and the NIHR (Cambridge Biomedical Research Centre at the Cambridge University Hospitals NHS Foundation Trust). The views expressed are those of the authors and not necessarily those of the NHS, the NIHR or the Department of Health and Social Care. A complete list of the investigators and contributors to the INTERVAL trial has previously been provided (1). The academic coordinating centre would like to thank blood donor centre staff and blood donors for participating in the INTERVAL trial. The Genotype-Tissue Expression (GTEx) project was supported by the Common Fund of the Office of the Director of the National Institutes of Health (NIH). Additional funds from the National Cancer Institute; National Human Genome Research Institute (NHGRI); National Heart, Lung, and Blood Institute; National Institute on Drug Abuse; National Institute of Mental Health; and National Institute of Neurological Disorders and Stroke. Donors were enrolled at Biospecimen Source Sites funded by Leidos Biomedical, Inc. (Leidos) subcontracts to the National Disease Research Interchange (10XS170) and Roswell Park Cancer Institute (10XS171). The Laboratory, Data Analysis and Coordinating Center (LDACC) was funded through a contract (HHSN268201000029C) to The Broad Institute, Inc. Biorepository operations were funded through a Leidos subcontract to Van Andel Institute (10ST1035). Additional data repository and project management provided by Leidos (HHSN261200800001E). The authors would like to thank the staff at the IUCPQ Biobank of the Quebec Respiratory Heath research Network for their valuable assistance with the lung eQTL dataset at Laval University.

